# Face-brain correlates as potential sex-specific biomarkers for schizophrenia and bipolar disorder

**DOI:** 10.1101/2024.01.23.24301590

**Authors:** Noemí Hostalet, Alejandro González, Rubèn Gonzàlez-Colom, Pilar Salgado-Pineda, Erick J Canales-Rodríguez, Candibel Aguirre, Amalia Guerrero-Pedraza, María Llanos-Torres, Raymond Salvador, Edith Pomarol-Clotet, Xavier Sevillano, Neus Martínez-Abadías, Mar Fatjó-Vilas

## Abstract

Given the shared ectodermal origin and integrated development of the face and the brain, facial biomarkers emerge as potential candidates to assess vulnerability for disorders in which neurodevelopment is compromised, such as Schizophrenia (SZ) and bipolar disorder (BD).

The sample comprised 188 individuals (67 SZ patients, 46 BD patients and 75 healthy controls (HC)). Using a landmark-based approach on 3D facial reconstructions, we quantified global and local facial shape differences between SZ/BD patients and HC using geometric morphometrics. We assessed correlations between facial and brain cortical measures. The analyses were performed separately by sex.

Diagnosis explained 4.1% - 5.9% of global facial shape variance in males and females with SZ, and 4.5% - 4.1% in BD. Regarding local facial shape, we detected 43.2% of significantly different distances in males and 47.4% in females with SZ compared to HC, whereas in BD the percentages decreased to 35.8% and 26.8%, respectively. We detected that brain area and volume significantly explained 2.2% and 2% of facial shape variance in the male SZ - HC sample.

Our results support facial shape as a neurodevelopmental marker for SZ and BD and reveal sex-specific pathophysiological mechanisms modulating the interplay between the brain and the face.

## 1. INTRODUCTION

Schizophrenia (SZ) and bipolar disorder (BD) are severe, chronic and disabling psychiatric disorders that affect approximately 65 million people worldwide and are considered among the top leading causes of years lived with disability (GBD 2019 Mental Disorders Collaborators, 2022).

The presence of a heterogeneous combination of symptoms that overlap in these disorders (Karantonis et al., 2020; Liu et al., 2020) combined with the diverse trajectory and response to treatment among patients, has hindered our understanding of the etiology of SZ and BD. This limitation has impeded the identification of biologically informed markers that could enable earlier and more accurate diagnostics. Growing evidence suggests that both environmental and genetic factors can impact brain development and maturation processes, and deviations from typical developmental trajectories may contribute to an elevated risk of various psychiatric disorders (Birnbaum and Weinberger, 2017; Owen and O’Donovan, 2017; Weinberger, 1995; Weinberger and Levitt, 2011). In this regard, the neurodevelopmental model is widely accepted as the main etiological model for SZ (Weinberger and Levitt, 2011), whereas in BD, the neurodevelopmental background is considered particularly prominent in individuals with an earlier onset, exhibiting psychotic symptoms and experiencing premorbid cognitive impairments (Kloiber et al., 2020).

Structural brain phenotypes have been broadly characterized through neuroimaging approaches as direct markers of neurodevelopmental trajectories (Ching et al., 2022; Howes et al., 2023). Although significant neuroanatomical differences have been identified between patients with SZ/BD and healthy individuals (Hibar et al., 2018; Jauhar et al., 2022; van Erp et al., 2018), these markers show heterogeneity and still cannot be reliably used in a clinical scenario.

Minor Physical Anomalies (MPAs) have also been widely studied in the context of the neurodevelopmental hypothesis. MPAs are slight dysmorphic features, without functional or cosmetic consequences, that are undistinguishable to the untrained eye and commonly present in the craniofacial region (facial dysmorphologies) and limbs due to developmental deviations.

MPAs are thus recognized as indirect markers of neurodevelopment according to the shared embryonic ectoderm origin of the epidermis and the central nervous system and the physical and genetic mechanisms underlying their integrated development (Marcucio, Ralph Hallgrimsson and Young, 2015; Naqvi et al., 2021; Waddington et al., 1999). Furthermore, the reported prevalence of MPAs is higher in patients with SZ and BD than in healthy individuals (Akabaliev et al., 2014; Bora, 2022; Gourion et al., 2004; Lane et al., 1997) and is considered a risk factor for these disorders (Radua et al., 2018), reinforcing their neurodevelopmental etiology.

Facial dysmorphologies have been classically assessed by linear or angular measurements and applying categorical scores (Akabaliev et al., 2014; Lane et al., 1997). The main disadvantage of these classical methodologies is their lack of precision and limited potential to capture subtle dysmorphologies in complex 3D anatomies. Therefore, in the search for higher accuracy, geometric morphometrics emerges as a more powerful approach to capture the subtle, yet significant, facial dysmorphologies associated with neurodevelopmental disorders. The few studies based on geometric morphometrics currently available have revealed significant facial shape differences between SZ/BD patients and healthy individuals (Buckley et al., 2005; Hennessy et al., 2010, 2007, 2004; Henriksson et al., 2006; Katina et al., 2020).

Despite the structural brain alterations and facial dysmorphologies detected in SZ and BD, few studies have explored the link between facial and brain dysmorphologies. Facial MPAs assessed by classical scales have been associated with grey matter changes in several brain areas (prefrontal cortex, precuneus basal ganglia, thalamus and lingual gyrus) of patients with first-episode psychosis (Dean et al., 2006) and lateral ventricular enlargement in childhood or adolescent-onset SZ patients (Hata et al., 2003). However, such associations were not detected by other studies (Kelly et al., 2005; O’Callaghan et al., 1995).

This study aimed to further evaluate facial dysmorphologies and their cortical brain measures correlates both in SZ and BD, by combining geometric morphometrics and neuroimaging approaches, to better understand the neurodevelopmental load of these disorders. Considering the existence of facial sexual dimorphism (Evison et al., 2010; Hennessy et al., 2002) together with the underlying sex-dependent mechanisms that influence the neurodevelopmental processes and produce structural brain disparities between males and females (Barrett and Lessing, 2021; Bethlehem et al., 2022; Diáz-Caneja et al., 2021; Sandman et al., 2013), it becomes imperative to embrace a sex-specific perspective in the evaluation of brain-face correlates. Furthermore, differences observed in the prevalence, age of onset and progression across sexes (Abel et al., 2010) emphasize the importance of this sex-specific approach. Therefore, we analyzed separately males and females to: i) assess global and local facial shape differences between patients with SZ and BD as compared with healthy individuals; and ii) evaluate the relationship between global facial shape and cortical brain measures. We expected to reveal significant differences in facial shape between groups and associations with brain cortical measures, adding evidence to the understanding of common developmental roots between the face and the brain in these disorders.

## 2. METHODS AND MATERIALS

### 2.1. Sample

The sample comprised 188 subjects, including 67 patients with SZ, 46 patients with BD and 75 healthy controls (HC). Patients were recruited at Benito Menni and Mare de Déu de la Mercè Hospitals, both of which are part of *Germanes Hospitalàries* centers in the province of Barcelona. They fulfilled the Diagnostic and Statistical Manual of Mental Disorders (4th edition, text revision) (DSM-IV-TR) criteria for SZ or BD according to clinical interview with psychiatrists. HC were recruited through public advertising in the area of Barcelona and did not have a current or antecedent history or first-degree family history of psychiatric disorders. In addition, both patients and HC met additional inclusion criteria, including European ancestry; age between 18 and 65 years old; right-handedness; and estimated intelligence quotient (IQ) > 70, as assessed using the *Test de acentuación de palabras* (TAP) (Gomar et al., 2011), an adapted Spanish version of the National Adult Reading Test (NART) (Nelson and Willison, 1991). Exclusion criteria for both groups included drug or alcohol abuse and a history of neurological damage. All participants provided written consent after being informed about the study procedures and implications. The study was performed following the guidelines of the institutions involved and was approved by the local ethics committee of the centers (PR-2023-21 and IRB00003099-CER122317). All procedures were carried out according to the Declaration of Helsinki.

Based on the clinical diagnosis, the global sample was subdivided into two subsamples (SZ-HC and BD-HC), in which patients of each disorder were matched for age, sex and estimated IQ with individuals from the HC group. All subsequent analyses were separately performed in each subsample. The sample characteristics are presented in Table 1.

**Table 1.** Sample description. The data presented in this table are categorized based on the groups used in the analyses. Healthy controls (HC) were matched with the corresponding patient groups (diagnosed with schizophrenia (SZ) or bipolar disorder (BD)) in terms of age, sex, and estimated IQ.

|  | **SZ – HC sample (n = 134)** |  | **BD – HC sample (n = 92)** |  |
| --- | --- | --- | --- | --- |
| **Diagnosis** | **SZ** | **HC** | **BD** | **HC** |
| **n (male/female)** | 67 (41/26) | 67 (41/26) | 46 (23/23) | 46 (23/23) |
| **Mean age (s.d.) [age range]** | 39.81 (10.19) [21-60] | 38.27 (8.99) [20 - 63] | 41.17 (9.88) [22-61] | 40.48 (9.14) [21-57] |
| **Estimated IQ** | 101 (8.6) [77-114] | 102.07(7.92) [83-116] | 102.93 (7.73) [81-114] | 102.33 (8.35) [83-116] |

### 2.2. Magnetic resonance images acquisition

High-resolution T1 weighted (T1w) head magnetic-resonance images (MRI) were obtained for all participants at Hospital Sant Joan de Déu (Esplugues de Llobregat, Spain). The neuroimaging protocol was conducted in a 1.5T GE Sigma scanner with the following acquisition parameters: matrix size 512 × 512; 180 contiguous axial slices; voxel resolution 0.47 × 0.47 × 1 mm3; echo (TE), repetition (TR) and inversion (TI) times, (TE/TR/TI) = 3.93ms/2000ms/710ms, respectively; and flip angle 15. These images were used to obtain both facial reconstructions and brain cortical measures.

### 2.3. Facial shape

#### 2.3.1. 3D facial reconstructions and morphometric phenotyping

3D facial reconstructions were generated from the MRIs using the 3D analysis software for scientific data Amira 2019.2 (Thermo Fisher Scientific, Waltham, MA, USA). Facial isosurfaces were created by selecting a grey-scale threshold value that optimized skin segmentation for each participant and facial region.

With the same software, the 3D coordinates of 20 anatomical facial landmarks were manually recorded on the facial surface by a trained researcher (RG) (Figure 1). This landmark selection was adapted from the original set of 26 anatomical landmarks previously used to assess the facial dysmorphologies associated with SZ (24). It included homologous landmarks that could be reliably located on the facial structures of all individuals in the sample. Landmark definitions are provided in Supplementary Table S1.

**Figure 1.**
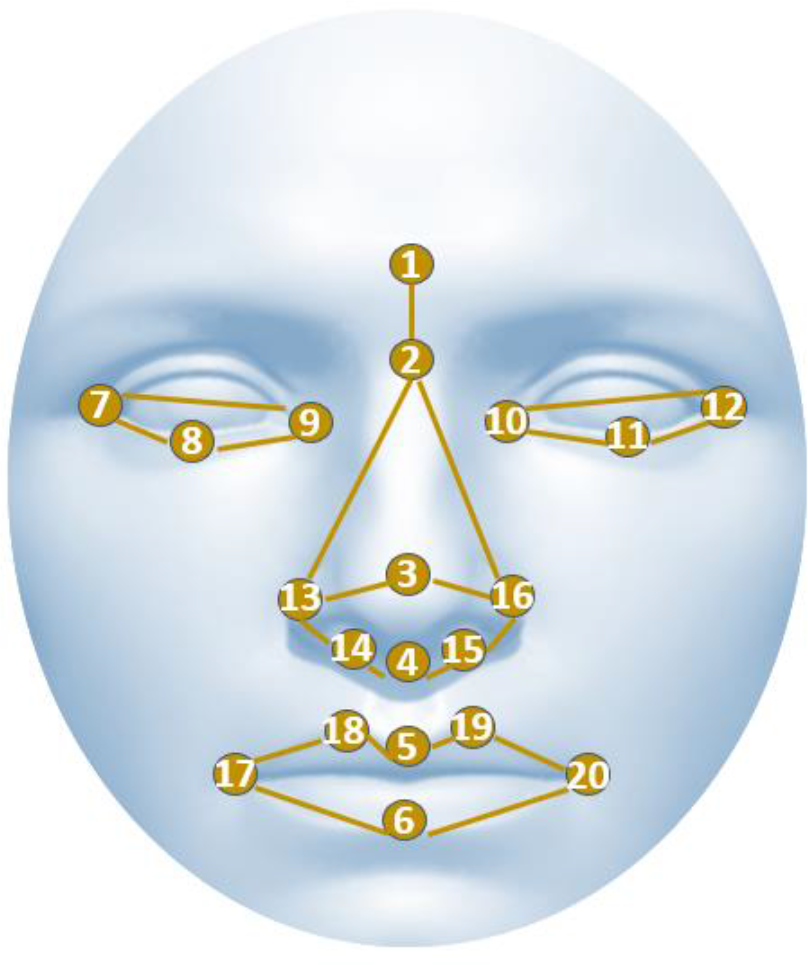
3D facial reconstruction showing the location of the analyzed 20 anatomical landmarks (round numerical symbols). Landmark definitions are provided in Supplementary Materials 1, Table S1.

#### 2.3.2. Facial shape morphometric analyses

Geometric Morphometrics (GM) was used to statistically compare facial shape variation among groups of patients and HC. GM is a sophisticated body of robust statistical tools developed for measuring and comparing 3D shapes with increased precision and efficiency (Bookstein, 1992; Dryden and Mardia, 2016; Mitteroecker and Schaefer, 2022) . In contrast to other traditional morphometrics techniques, the main advantage of GM is that the 3D shape and geometry are preserved throughout the analysis, allowing the visualization of the results as shape changes (Mitteroecker and Gunz, 2009).

To place the landmark configurations from all the individuals in a common morphospace, a full Generalized Procrustes Analysis (GPA) was performed with MorphoJ (Klingenberg, 2011). This procedure removes the influence of size and adopts a single orientation for all the individuals by shifting the landmark configurations to a common position, scaling them to a standard size and rotating them until achieving a least-squares fit of corresponding landmarks. The resulting Procrustes coordinates represent the facial shape of each individual.

As an estimated measure of facial size, centroid size (CS) was computed as the square root of the summed squared distances between each landmark coordinate and the centroid of the configuration. The log-transformed CS (log (CS)) was used for the analyses.

The Procrustes Distance (PD), which measures the similarity or dissimilarity between shapes based on their Procrustes shape, was used to assess the global differences in facial shape between groups (Mitteroecker and Gunz, 2009). To evaluate these differences, PD values were calculated between the average shapes of each group using 10,000 permutations. Global facial shape variation within each sample was then assessed by performing a Principal Components Analysis (PCA) (Hallgrimsson et al., 2015).

From the Procrustes shape coordinates, we also quantified the relative amount of facial shape variation attributable to the diagnostic group and other variables that could be relevant for facial shape variation, such as age and size (Kesterke et al., 2016; Larson et al., 2018), by applying a Procrustes ANOVA with 1,000 permutations using the function ProcD.lm implemented in Geomorph R package (Adams et al., 2019).

To control for the potential confounding effects of age and size on facial shape variance in the local facial shape analyses, a multivariate regression of the Procrustes coordinates on age and size was first performed (Klingenberg, 2016), and the resulting regression residuals were used to conduct the following morphometric analyses.

To precisely localize the specific facial traits associated with SZ and BD, an Euclidean Distance Matrix Analysis (EDMA) was applied using MATLAB v.9.13.0 (The MathWorks Inc, 2022). EDMA represented each subject as a matrix of 190 linear distances between all possible pairs of anatomical landmarks and pinpointed precisely which linear measurements differed between groups of patients and controls through pairwise comparisons based on confidence interval testing (α = 0.10) (Lele and Richtsmeier, 2001). The percentage of significantly different interlandmark distances between affected (SZ/BD) and unaffected (HC) individuals was considered as the Facial Difference Score (FDS) (Starbuck et al., 2021). To reveal the unique morphological patterns of variation associated with each pairwise contrast, the 20 most significant local shape differences were plotted upon facial figures, identifying the 10 largest and shortest distances in patients compared with HC.

Finally, we performed a series of EDMA simulations based on artificial samples that were derived from the parametric distributions of the original samples. This approach statistically assessed whether the percentage of different distances between patients and HC (FDS) was statistically different from the percentage obtained from a random distribution of HC. In each simulation, a group composed exclusively of random HC individuals was compared to a mixed group composed of random HC and patients (running 200 iterations). In the first simulation, the mixed group was composed of 100% HC. Therefore, the comparison was performed exclusively among HC. The process was conducted 6 times, increasing by 20% the percentage of patients in the mixed group in each simulation, reaching 100% in the last simulation. The p-value was calculated as the number of iterations in which the EDMA result obtained in the first simulation (in which the two groups only included HC) was larger than the EDMA result based on the complete sample of patients and HC (FDS), divided by the total number of EDMA iterations (200). A detailed explanation of this method is provided in Supplementary Materials 2.

### 2.4. Brain cortical measures

#### 2.4.1. Brain cortical measures phenotyping

Brain cortical data were obtained from the T1w MRIs applying the FreeSurfer protocol (http://surfer.nmr.mgh.harvard.edu/). Image pre-processing included motion correction, removal of non-brain tissue, automated Talairach transformation, tessellation of the grey and white matter boundaries and surface deformation (Fischl et al., 2004). This method uses both intensity and continuity information from the entire 3D images in the segmentation and deformation procedures to produce vertex-wise representations of cortical surface area (SA), cortical thickness (CT) and cortical volume (CV). FreeSurfer automatically segments the brain cortex into 34 cortical grey matter regions per hemisphere based on the Desikan–Killiany atlas, resulting in a total of 68 measures of SA, 68 measures of CT and 68 measures of CV (Desikan et al., 2006) (Supplementary Materials 1, Table S2). All the images included in this study complied with the standardized quality control protocols from the ENIGMA consortium (http://enigma.ini.usc.edu/protocols/imaging-protocols).

#### 2.4.2. Brain cortical measures analyses

To reduce the high dimensionality of cortical brain data, a PCA was performed (Fernandez-Cabello et al., 2022). Considering the different developmental trajectories of SA and CT and their interaction in CV (Norbom et al., 2021; Wierenga et al., 2014; Winkler et al., 2010), specific analyses were performed separately for each type of cortical measure. First, we performed linear regressions to control for age and intracranial volume (ICV) effects in SZ – HC /BD – HC groups separately by sex. The regression residuals were used then as the input for PCA.

### 2.5. Association analysis of brain cortical measures with facial shape

Procrustes ANOVA models were built to assess the facial shape variance attributable to the PC1 of SA, CT and CV. In the initial analysis (reduced model), the diagnosis was excluded from the model, whereas it was included in a second model (full model). A log-likelihood ratio test was applied to assess which model better fitted the facial shape data.

## 3. RESULTS

### 3.1. Facial shape morphometric analyses

Procrustes distances (PD) between groups indicated that the global facial shape of patients and HC was significantly different in both sexes when comparing SZ – HC (males: PD = 0.0313; females: PD = 0.038; p < 0.0001) and BD – HC (males: PD = 0.034; p = 0.001; females: PD = 0.028; p = 0.02). In both SZ and BD diagnoses, while statistically significant, the differences observed between patients and HC were subtle, as evidenced by the extensive overlap of both diagnostic groups in all the PCAs (Figure 2).

**Figure 2.**
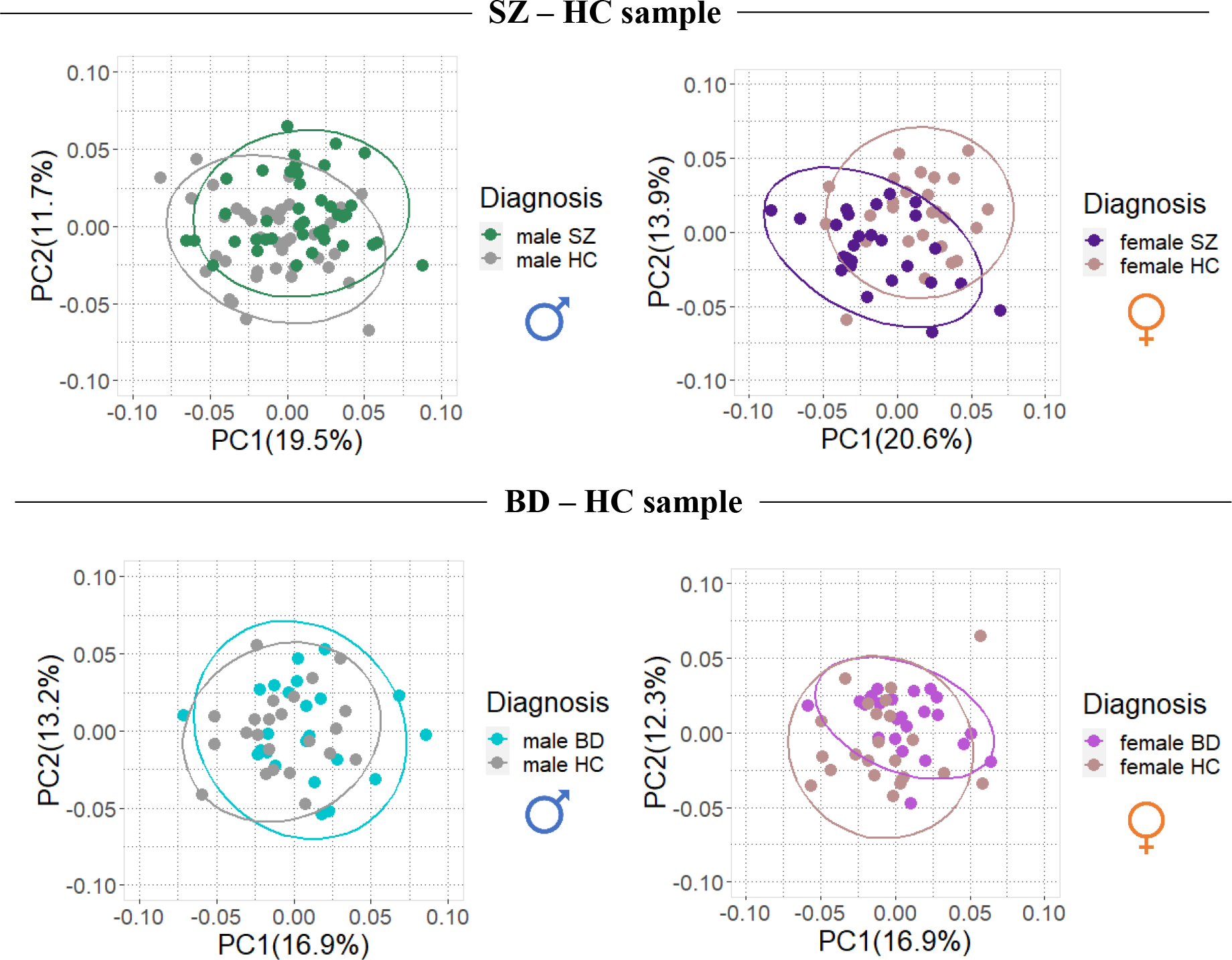
Principal Component Analyses corresponding to SZ – HC males (A); SZ – HC females (B); BD – HC males (C); and BD – HC females (D). Scatterplots of the PC1 and PC2 axes with the corresponding percentage of total morphological variance explained are displayed along each axis.

Procrustes ANOVA analyses reinforced the previous results, quantifying a significant effect of diagnosis on facial shape variance in both psychiatric disorders. In the SZ-HC sample, diagnosis explained 4.2% of facial shape variance in males (F = 3.734, p = 0.001) and 5.9% in females (F = 3.294, p = 0.001). In the BD – HC sample, diagnosis explained 4.5% of variance in males (F = 2.23, p = 0.006) and 4.1% in females (F = 1.95, p = 0.016). Full models are shown in Supplementary Materials 1, Tables S3 – S4.

When analyzing local facial shape differences between patients and controls, EDMA revealed that 43.2% of the 190 facial distances between SZ – HC males were significantly different, whereas for SZ – HC females, the percentage increased to 47.4%. The percentage of significantly different facial distances was lower in the comparison between BD patients and HC. The percentage of significant differences in males was 35.8%, whereas in females, 26.8% of facial distances were significantly different. A description of all the significant interlandmark distances is provided in Supplementary Materials 1, Table S5.

Through simulations, it was validated that, apart from the comparison of BD – HC females (p = 0.055), the facial differences identified between patients and HC were not coincidental but significantly associated with the diagnosis (p < 0.05) in all other comparisons.

When considering the top 10 significantly shorter and larger distances for patients (Table 2), the following morphological patterns emerged. In SZ, differences in patients as compared to HC were mainly localized in the eyes and mouth regions. Specifically, patients showed smaller and more separate eyes, smaller mouth, and an increased distance between the nose and mouth, as compared with HC, but showing a different pattern in males and females (Figure 3). In BD (Figure 4), male patients also showed smaller distances in the mouth region, but in this group, the main differences were localized in the distances between the mouth and the nose and between the nose and the eyes. In this case, patients showed larger distances in the upper face (between nose and eyes, and between the extremes of the eyes) and larger distances on the lower part of the face (between mouth and nose).

**Figure 3.**
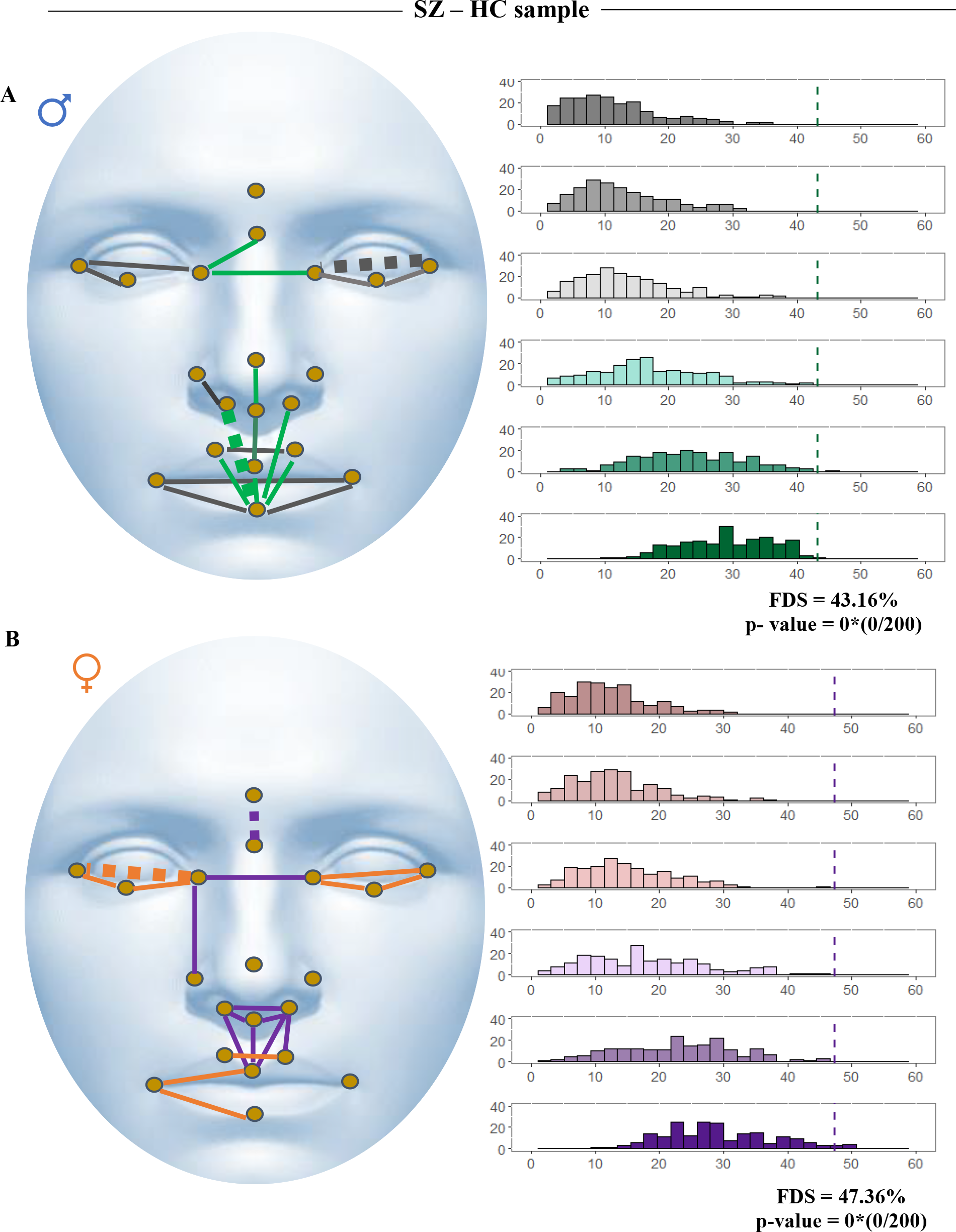
Representation of the 20 most significantly different distances from Euclidean Distance Matrix Analysis for SZ – HC males (A) and SZ – HC females (B) (left). Shorter distances for SZ patients are represented in grey (males) and orange (females). Larger distances for SZ patients are represented in green (males) and violet (females). Dashed lines represent the largest and shortest distances in SZ patients. Histograms represent the EDMA results for each simulation (right). An increasing percentage of different distances is obtained when more patients are added to the mixed group. The dashed line shows the FDS obtained with the original sample of HC and patients. *Statistically significant p-value.

**Figure 4.**
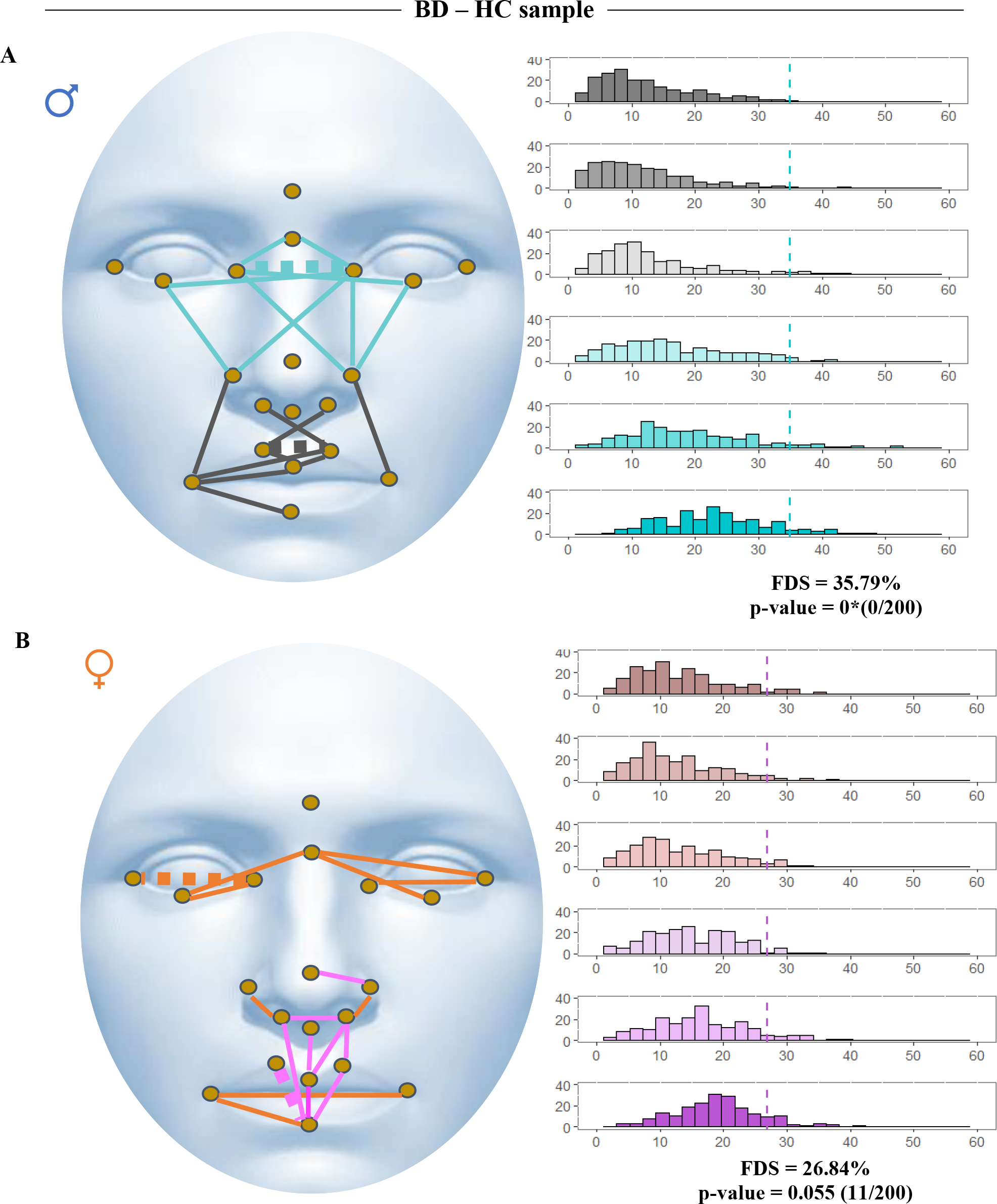
Representation of the 20 most significantly different distances from Euclidean Distance Matrix Analysis for BD – HC males (A) and BD – HC females (B) (left). Shorter distances for BD patients are represented in grey (males) and orange (females). Larger distances for BD patients are represented in turquoise (males) and pink (females). Dashed lines represent the largest and shortest distances in BD patients. Histograms represent the EDMA results for each simulation (right). An increasing percentage of different distances is obtained when more patients are added to the mixed group only in males since females did not achieve statistical significance. The dashed line shows the FDS obtained with the original sample of HC and patients. *Statistically significant p-value.

**Table 2.** Ranges of relative change in the top 10 distances that were significantly longer or shorter in patients compared to HC. A detailed list of all the distances showing significant differences between groups is provided in Supplementary Materials 1, Table S5.

|  |  | Longer distances in patients | Shorter distances in patients |
| --- | --- | --- | --- |
| SZ | males | 12.3% - 5.1% | 7.7% - 4.4% |
|  | females | 11.8% - 4.7% | 11% - 6.9% |
| BD | males | 7.2% - 4.2% | 19.4% - 5.3% |
|  | females | 10.2% - 5.4% | 9.5% - 3.7% |

### 3.2. Brain cortical measures analyses

The total variance explained by the PC1 of each PCA ranged from 17.71% to 38.77%. Since the variance explained by PC2 considerably decreased (Supplementary Materials 1, Table S6 –S17), and variables from all the cortical regions contributed to explain the morphological variance captured by PC1s (Supplementary materials 1, Table S6– Table S17), only PC1s were retained for further analyses. Area PC1, Thickness PC1 and Volume PC1 were thus considered as our global cortical brain variables in the subsequent statistical models.

### 3.3. Association analysis of brain cortical measures with facial shape

We only detected a significant association between brain cortical measures and facial shape in the SZ–HC male sample. Procrustes ANOVA showed that after controlling for age and size covariates, Area PC1 and Volume PC1 had a significant effect on facial shape variance (F = 2.76, p = 0.003; and F = 2.45, p = 0.008), explaining respectively 3.1% and 2.6% of facial shape variance. When the diagnosis was added to the model, the effect was maintained and Area PC1 (F = 2.06, p = 0.022) and Volume PC1 (F = 1.89, p = 0.03) significantly explained 2.2% and 2% of facial shape variance (Figure 5). Thickness PC1 did not significantly affect facial shape in any model (p > 0.05).

**Figure 5.**
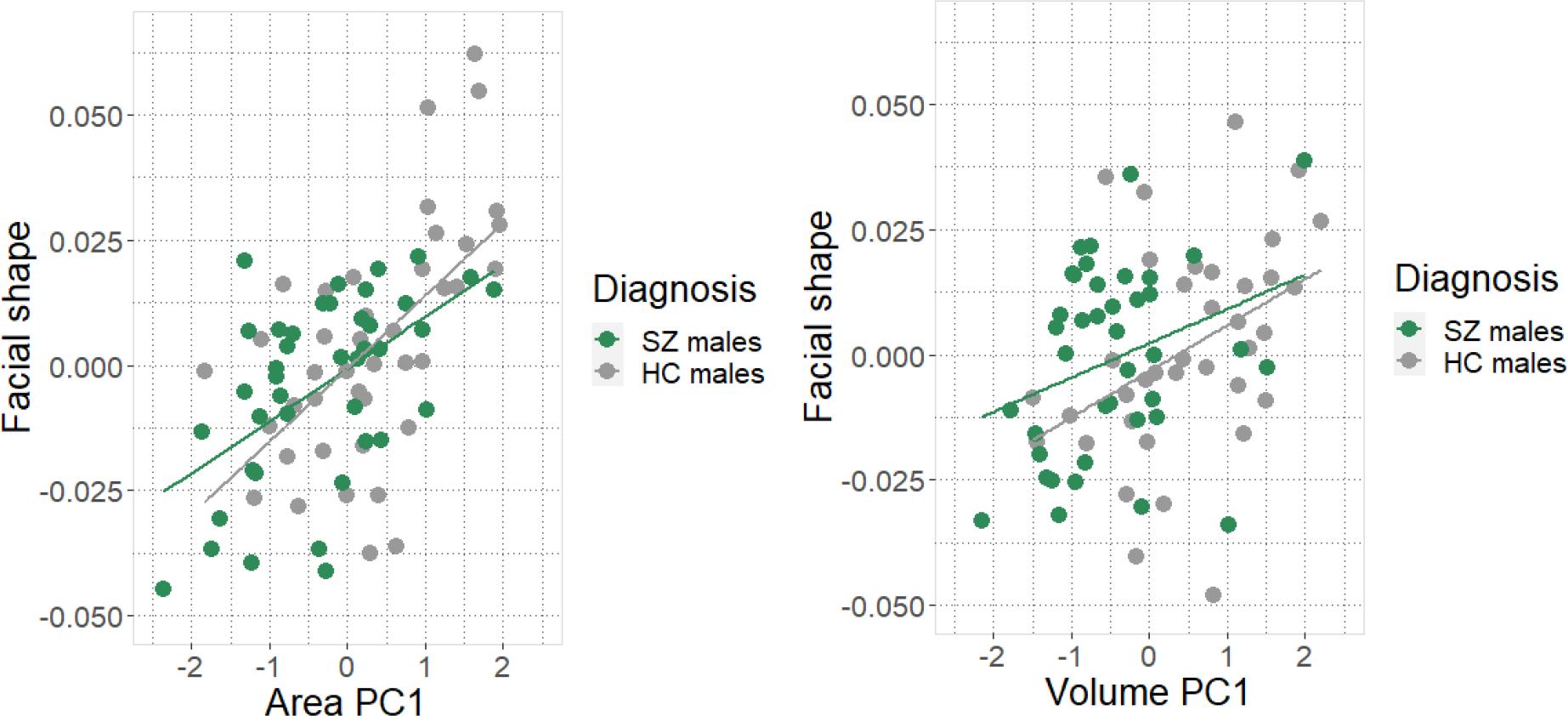
Regression scores scatter plots from Procrustes ANOVA showing the association dependent on diagnosis of Area PC1 (left) and Volume PC1 (right) with facial shape.

Despite the decrease of variance explained by SA and CV when introducing the diagnosis (full models), the log-likelihood test revealed that the full model had better fitness than the reduced one in the case of Area PC1 (F = 2.94, R^2^= 3.3%, p = 0.001) and Volume PC1 (F = 1.82, R^2^= 3.1%, p = 0.001).

## 4. DISCUSSION

In this study, we used neuroimaging and morphometric techniques to evaluate face-brain correlates along the health-psychosis continuum. Following a sex-specific perspective, we explored face/brain associations within SZ and BD and their combined potential role as biomarkers.

First, we reported a subtle but significant effect of diagnosis on global facial morphology in both SZ and BD patients. These findings align with previous research demonstrating a connection between SZ and BD diagnoses, and alterations in overall facial shape among males (Buckley et al., 2005; Hennessy et al., 2010, 2007, 2004), females (Hennessy et al., 2007) and sex-pooled samples (Buckley et al., 2005). This differs from other studies that did not detect any SZ diagnosis impact in females (Hennessy et al., 2004) or when analyzing combined samples (Katina et al., 2020). Discrepancies across studies may stem from variations in sample sizes, male-to-female ratios, and the lack of control over confounding factors such as size and age, which can influence global facial shape.

Second, our in-depth analyses of local facial shape differences between SZ/BD patients and healthy individuals consistently extended the global facial shape results. The highest percentage of significantly different distances between patients and HC was obtained in SZ, whereas in BD, the number and magnitude of relative facial differences were lower. Our simulation approach confirmed that the morphological differences between patients and healthy individuals were not due to chance in any group, except for BD females, which could indicate a lower effect of diagnosis on facial shape in this group. However, the reduced sample size and the restrictiveness of the method could also explain this result.

Regarding the characteristic local morphology of SZ patients as compared to HC, previous studies have consistently noted the presence of hypertelorism in both male and female SZ patients by using geometric morphometric methods (Henriksson et al., 2006) or classical linear measurements (Akabaliev et al., 2011; Lawrie et al., 2001). In addition, our results revealed sex-specific morphological differences. Females with SZ exhibited a wider nose base distance than female controls, a pattern that was not detected in males. Concerning the local morphology of BD male patients as compared to HC, our study identified longer distances on the upper midface and shorter distances on the lower midface among patients. These findings align with the characteristic facial morphology previously reported in BD male patients, exhibiting elongated noses and narrower mouths (Hennessy et al., 2007).

To our knowledge, this is the first study assessing local facial differences in SZ and BD applying EDMA. Therefore, our results cannot be directly compared with any other study and further studies in new samples should be performed to validate them. However, our data underscores the interest of the precise characterization of sex-specific morphological patterns associated with each disorder. This knowledge is crucial to establish the biological and morphological correlates the pathophysiology of SZ and BD.

While further studies with larger samples are needed to validate our results and broaden the evidence, our findings highlight global and local facial shape differences as potential neurodevelopmental markers associated with SZ and BD. Our study adds robustness to previous data by matching patients and healthy individuals for age, estimated IQ and sex; and controlling the potential confounding effect of size and age. Moreover, the use of advanced geometric morphometric tools has allowed us to detect facial differences previously described, as well as to provide for the first time an accurate description of the characteristic morphology of SZ and BD based on the whole set of 190 interlandmark distances. Therefore, our methods replicate and broaden the differences already found by classical methods.

Regarding the study of facial shape and brain correlates, our study adds novelty beyond the combination of classical MPAs assessments with volumetric voxel-based data of previous studies (Dean et al., 2006; Hata et al., 2003; Kelly et al., 2005; O’Callaghan et al., 1995). By using morphometric data and cortical measures tightly related to neurodevelopment (Patel et al., 2022), we reported a significant association of global facial shape with SA and CV in the SZ – HC males sample, although modest in terms of variance explained. Since the CV is the product of SA and CT, the significant association of global facial shape with CV might be related to the significant result obtained in global facial shape and SA association.

Differences in the association of SA and CT with global facial shape might be explained by the different ontogenies of SA and CT (Norbom et al., 2021; Wierenga et al., 2014; Winkler et al., 2010), as well as by the different genetic influences in each measure, as shown by heritability estimates (SA: h^2^= 0.78; CT: h^2^=0.29) (Jha et al., 2018). Furthermore, the genetic regulatory processes related to SA and CT are different, being related to gene regulatory activity in neural progenitor cells during fetal development for SA, while those for CT occur after mid-fetal development (Grasby et al., 2020).

The association between global facial shape and brain cortical measures, limited to SZ – HC males, might indicate the presence of sex-specific pathophysiological mechanisms that influence the dialogue between the brain and the face during neurodevelopment. On the other hand, the absence of significant results in the association between facial shape and cortical measures in the other samples might be attributed to an insufficient statistical power to capture more nuanced relationships, probably due to smaller sample sizes.

To strengthen these findings, future studies should increase the sample sizes of patients and HC, and ensure a more homogeneous representation of developmental stages, including in the analyses individuals within narrower and more closely matched age ranges that minimize the significant influence of age on facial shape. While the total sample size analyzed in this study was relatively larger in comparison to similar studies, our aim of exploring sex differences involved the subdivision of the sample, resulting in reduced subsets for each sex and diagnostic group. Despite this trade-off, our results underscore that adopting a sex-specific perspective in neurodevelopment research can provide valuable insights and reveal sex-specific differences in psychotic disorders that otherwise remain unveiled and may lead to misleading results.

The significant facial shape differences detected between patients and healthy individuals by our quantitative analyses highlight the potential of facial biomarkers in SZ and BD. Facial biomarkers may be used in the future to improve the clinical management of patients with these disorders, in particular at the early stages of psychotic symptoms manifestation, when it is challenging to provide a first diagnosis. However, the facial differences reported here were very subtle and undetectable by the naked eye, discarding their value as a screening tool in the general population.

Since facial biomarkers may have a potential application in biomedical context but also entail ethical issues and social implications, it is crucial to ensure that these findings are only used with scientific or medical purposes, protecting the highly sensitive nature of the data.

Overall, our results highlight facial shape biomarkers as promising candidates to assess biological risk for SZ and BD and reveal sex-specific pathophysiological mechanisms modulating the interplay between the brain and the face during neurodevelopment in SZ. Our results encourage further research to establish face-brain correlates as biomarkers for these disorders, performing even more accurate morphometric analyses integrated with genetic data. This approach would facilitate the development of a composite biomarker that combines brain-face phenomic data and genomic data, ultimately enhancing its potential for assessing vulnerability to SZ and BD.

## Data Availability

All data produced in the present study are available upon reasonable request to the authors

## Acknowledgements

This study received funding from: i) Agencia Española de Investigación (PID2020-113609RB- C21 /AEI/10.13039/501100011033); ii) Instituto de Salud Carlos III (ISCIII) through the project PI20/01002 and the contracts FI21/00093 to NH and CP20/00072 to MF-V (co-funded by European Regional Development Fund (ERDF)/European Social Fund “Investing in your future”); iii) the Comissionat per a Universitats i Recerca del DIUE of the Generalitat de Catalunya (Agència de Gestio d’Ajuts Universitaris i de Recerca (AGAUR: 2021SGR01475, 2021SGR01396, and 2021SGR00706).

## References

Abel, K.M., Drake, R., Goldstein, J.M., 2010. Sex differences in schizophrenia. International Review of Psychiatry 22, 417–428. 10.3109/09540261.2010.515205

Adams, D.C., Collyer, M.L., Kahliotzopolou, A., 2019. Geomorph: Software for geometric morphometric analyses. R package version 3.1.0.

Akabaliev, V.H., Sivkov, S.T., Mantarkov, M.J., Ahmed-Popova, F.M., 2011. Biomarker profile of minor physical anomalies in schizophrenia patients. Folia Med (Plovdiv) 53, 45–51. 10.2478/v10153-011-0056-z

Akabaliev, V.H., Sivkov, S.T., Mantarkov, M.Y., 2014. Minor physical anomalies in schizophrenia and bipolar I disorder and the neurodevelopmental continuum of psychosis. Bipolar Disord 16, 633–641. 10.1111/bdi.12211

Barrett, E.S., Lessing, J., 2021. Sex-Specific Impacts of Prenatal Stress, in: Wazana, A., Székely, E., Oberlander, T.F. (Eds.), Prenatal Stress and Child Development. Springer International Publishing, Cham, pp. 243–277. 10.1007/978-3-030-60159-1_10

Bethlehem, R.A.I., Seidlitz, J., White, S.R., Vogel, J.W., Anderson, K.M., Adamson, C., Adler, S., Alexopoulos, G.S., Anagnostou, E., Areces-Gonzalez, A., Astle, D.E., Auyeung, B., Ayub, M., Bae, J., Ball, G., Baron-Cohen, S., Beare, R., Bedford, S.A., Benegal, V., Beyer, F., Blangero, J., Blesa Cábez, M., Boardman, J.P., Borzage, M., Bosch-Bayard, J.F., Bourke, N., Calhoun, V.D., Chakravarty, M.M., Chen, C., Chertavian, C., Chetelat, G., Chong, Y.S., Cole, J.H., Corvin, A., Costantino, M., Courchesne, E., Crivello, F., Cropley, V.L., Crosbie, J., Crossley, N., Delarue, M., Delorme, R., Desrivieres, S., Devenyi, G.A., Di Biase, M.A., Dolan, R., Donald, K.A., Donohoe, G., Dunlop, K., Edwards, A.D., Elison, J.T., Ellis, C.T., Elman, J.A., Eyler, L., Fair, D.A., Feczko, E., Fletcher, P.C., Fonagy, P., Franz, C.E., Galan-Garcia, L., Gholipour, A., Giedd, J., Gilmore, J.H., Glahn, D.C., Goodyer, I.M., Grant, P.E., Groenewold, N.A., Gunning, F.M., Gur, R.E., Gur, R.C., Hammill, C.F., Hansson, O., Hedden, T., Heinz, A., Henson, R.N., Heuer, K., Hoare, J., Holla, B., Holmes, A.J., Holt, R., Huang, H., Im, K., Ipser, J., Jack, C.R., Jackowski, A.P., Jia, T., Johnson, K.A., Jones, P.B., Jones, D.T., Kahn, R.S., Karlsson, H., Karlsson, L., Kawashima, R., Kelley, E.A., Kern, S., Kim, K.W., Kitzbichler, M.G., Kremen, W.S., Lalonde, F., Landeau, B., Lee, S., Lerch, J., Lewis, J.D., Li, J., Liao, W., Liston, C., Lombardo, M. V., Lv, J., Lynch, C., Mallard, T.T., Marcelis, M., Markello, R.D., Mathias, S.R., Mazoyer, B., McGuire, P., Meaney, M.J., Mechelli, A., Medic, N., Misic, B., Morgan, S.E., Mothersill, D., Nigg, J., Ong, M.Q.W., Ortinau, C., Ossenkoppele, R., Ouyang, M., Palaniyappan, L., Paly, L., Pan, P.M., Pantelis, C., Park, M.M., Paus, T., Pausova, Z., Paz-Linares, D., Pichet Binette, A., Pierce, K., Qian, X., Qiu, J., Qiu, A., Raznahan, A., Rittman, T., Rodrigue, A., Rollins, C.K., Romero-Garcia, R., Ronan, L., Rosenberg, M.D., Rowitch, D.H., Salum, G.A., Satterthwaite, T.D., Schaare, H.L., Schachar, R.J., Schultz, A.P., Schumann, G., Schöll, M., Sharp, D., Shinohara, R.T., Skoog, I., Smyser, C.D., Sperling, R.A., Stein, D.J., Stolicyn, A., Suckling, J., Sullivan, G., Taki, Y., Thyreau, B., Toro, R., Traut, N., Tsvetanov, K.A., Turk-Browne, N.B., Tuulari, J.J., Tzourio, C., Vachon-Presseau, Valdes-Sosa, M.J., Valdes-Sosa, P.A., Valk, S.L., van Amelsvoort, T., Vandekar, S.N., Vasung, L., Victoria, L.W., Villeneuve, S., Villringer, A., Vértes, P.E., Wagstyl, K., Wang, Y.S., Warfield, S.K., Warrier, V., Westman, E., Westwater, M.L., Whalley, H.C., Witte, A. V., Yang, N., Yeo, B., Yun, H., Zalesky, A., Zar, H.J., Zettergren, A., Zhou, J.H., Ziauddeen, H., Zugman, A., Zuo, X.N., Rowe, C., Frisoni, G.B., Binette, A.P., Bullmore, E.T., Alexander-Bloch, A.F., 2022. Brain charts for the human lifespan. Nature 604, 525–533. 10.1038/s41586-022-04554-y

Birnbaum, R., Weinberger, D.R., 2017. Genetic insights into the neurodevelopmental origins of schizophrenia. Nat Rev Neurosci 18, 727–740. 10.1038/nrn.2017.125

Bookstein, F.L., 1992. Morphometric Tools for Landmark Data. Cambridge University Press. 10.1017/CBO9780511573064

Bora, E., 2022. Minor physical anomalies in bipolar disorder in comparison to healthy controls and schizophrenia: A systematic review and meta-analysis. European Neuropsychopharmacology 65, 4–11. 10.1016/j.euroneuro.2022.08.007

Buckley, P.F., Dean, D., Ph, D., Bookstein, F.L., Ph, D., Min, K., Singer, B., 2005. A Three-Dimensional Morphometric Study of Craniofacial Shape in Schizophrenia. American Journal of Psychiatry 162, 606–608.

Ching, C.R.K., Hibar, D.P., Gurholt, T.P., Nunes, A., Thomopoulos, S.I., Abé, C., Agartz, I., Brouwer, R.M., Cannon, D.M., de Zwarte, S.M.C., Eyler, L.T., Favre, P., Hajek, T., Haukvik, U.K., Houenou, J., Landén, M., Lett, T.A., McDonald, C., Nabulsi, L., Patel, Y., Pauling, M.E., Paus, T., Radua, J., Soeiro-de-Souza, M.G., Tronchin, G., van Haren, N.E.M., Vieta, E., Walter, H., Zeng, L.L., Alda, M., Almeida, J., Alnæs, D., Alonso-Lana, S., Altimus, C., Bauer, M., Baune, B.T., Bearden, C.E., Bellani, M., Benedetti, F., Berk, M., Bilderbeck, A.C., Blumberg, H.P., Bøen, E., Bollettini, I., del Mar Bonnin, C., Brambilla, P., Canales-Rodríguez, E.J., Caseras, X., Dandash, O., Dannlowski, U., Delvecchio, G., Díaz-Zuluaga, A.M., Dima, D., Duchesnay, É., Elvsåshagen, T., Fears, S.C., Frangou, S., Fullerton, J.M., Glahn, D.C., Goikolea, J.M., Green, M.J., Grotegerd, D., Gruber, O., Haarman, B.C.M., Henry, C., Howells, F.M., Ives-Deliperi, V., Jansen, A., Kircher, T.T.J., Knöchel, C., Kramer, B., Lafer, B., López-Jaramillo, C., Machado-Vieira, R., MacIntosh, B.J., Melloni, E.M.T., Mitchell, P.B., Nenadic, I., Nery, F., Nugent, A.C., Oertel, V., Ophoff, R.A., Ota, M., Overs, B.J., Pham, D.L., Phillips, M.L., Pineda-Zapata, J.A., Poletti, S., Polosan, M., Pomarol-Clotet, E., Pouchon, A., Quidé, Y., Rive, M.M., Roberts, G., Ruhe, H.G., Salvador, R., Sarró, S., Satterthwaite, T.D., Schene, A.H., Sim, K., Soares, J.C., Stäblein, M., Stein, D.J., Tamnes, C.K., Thomaidis, G. V., Upegui, C.V., Veltman, D.J., Wessa, M., Westlye, L.T., Whalley, H.C., Wolf, D.H., Wu, M.J., Yatham, L.N., Zarate, C.A., Thompson, P.M., Andreassen, O.A., 2022. What we learn about bipolar disorder from large-scale neuroimaging: Findings and future directions from the ENIGMA Bipolar Disorder Working Group. Hum Brain Mapp 43, 56–82. 10.1002/hbm.25098

Dean, K., Fearon, P., Morgan, K., Hutchinson, G., Orr, K., Chitnis, X., Suckling, J., Mallet, R., Leff, J., Jones, P.B., Murray, R.M., Dazzan, P., 2006. Grey matter correlates of minor physical anomalies in the ÆSOP first-episode psychosis study. British Journal of Psychiatry 189, 221–228. 10.1192/bjp.bp.105.016337

Desikan, R.S., Se, F., Fischl, B., Quinn, B.T., Dickerson, B.C., Blacker, D., Buckner, R.L., Dale, A.M., Maguire, R.P., Hyman, B.T., Albert, M.S., Killiany, R.J., 2006. An automated labeling system for subdividing the human cerebral cortex on MRI scans into gyral based regions of interest 31, 968–980. 10.1016/j.neuroimage.2006.01.021

Diáz-Caneja, C.M., Alloza, C., Gordaliza, P.M., Fernández-Pena, A., De Hoyos, L., Santonja, J., Buimer, E.E.L., Van Haren, N.E.M., Cahn, W., Arango, C., Kahn, R.S., Hulshoff Pol, H.E., Schnack, H.G., Janssen, J., 2021. Sex Differences in Lifespan Trajectories and Variability of Human Sulcal and Gyral Morphology. Cerebral Cortex 31, 5107–5120. 10.1093/cercor/bhab145

Dryden, I., Mardia, K., 2016. Statistical shape analysis, with applications in R: Second edition, Statistical Shape Analysis, with Applications in R: Second Edition. 10.1002/9781119072492

Evison, M., Dryden, I., Fieller, N., Mallett, X., Morecroft, L., Schofield, D., Bruegge, R.V., 2010. Key parameters of face shape variation in 3D in a large sample. J Forensic Sci 55, 159–162. 10.1111/j.1556-4029.2009.01213.x

Fernandez-Cabello, S., Alnæs, D., van der Meer, D., Dahl, A., Holm, M., Kjelkenes, R., Maximov, I.I., Norbom, L.B., Pedersen, M.L., Voldsbekk, I., Andreassen, O.A., Westlye, L.T., 2022. Associations between brain imaging and polygenic scores of mental health and educational attainment in children aged 9–11. Neuroimage 263, 119611. 10.1016/j.neuroimage.2022.119611

Fischl, B., Van Der Kouwe, A., Destrieux, C., Halgren, E., Ségonne, F., Salat, D.H., Busa, E., Seidman, L.J., Goldstein, J., Kennedy, D., Caviness, V., Makris, N., Rosen, B., Dale, A.M., 2004. Automatically Parcellating the Human Cerebral Cortex. Cerebral Cortex 14, 11–22. 10.1093/cercor/bhg087

GBD 2019 Mental Disorders Collaborators, 2022. Global, regional, and national burden of 12 mental disorders in 204 countries and territories, 1990&#x2013;2019: a systematic analysis for the Global Burden of Disease Study 2019. Lancet Psychiatry 9, 137–150. 10.1016/S2215-0366(21)00395-3

Gomar, J.J., Ortiz-Gil, J., McKenna, P.J., Salvador, R., Sans-Sansa, B., Sarró, S., Guerrero, A., Pomarol-Clotet, E., 2011. Validation of the Word Accentuation Test (TAP) as a means of estimating premorbid IQ in Spanish speakers. Schizophr Res 128, 175–176. 10.1016/j.schres.2010.11.016

Gourion, D., Bourdel, M., Bayle, F.J., Loo, H., 2004. Minor physical anomalies in patients with schizophrenia and their parents : prevalence and pattern of craniofacial abnormalities. Psychiatry Res 125, 21–28. 10.1016/j.psychres.2003.06.001

Grasby, K.L., Jahanshad, N., Painter, J.N., Colodro-Conde, L., Bralten, J., Hibar, D.P., Lind, P.A., Pizzagalli, F., Ching, C.R.K., McMahon, M.A.B., Shatokhina, N., Zsembik, L.C.P., Thomopoulos, S.I., Zhu, A.H., Strike, L.T., Agartz, I., Alhusaini, S., Almeida, M.A.A., Alnæs, D., Amlien, I.K., Andersson, M., Ard, T., Armstrong, N.J., Ashley-Koch, A., Atkins, J.R., Bernard, M., Brouwer, R.M., Buimer, E.E.L., Bülow, R., Bürger, C., Cannon, D.M., Chakravarty, M., Chen, Q., Cheung, J.W., Couvy-Duchesne, B., Dale, A.M., Dalvie, S., de Araujo, T.K., de Zubicaray, G.I., de Zwarte, S.M.C., den Braber, A., Doan, N.T., Dohm, K., Ehrlich, S., Engelbrecht, H.R., Erk, S., Fan, C.C., Fedko, I.O., Foley, S.F., Ford, J.M., Fukunaga, M., Garrett, M.E., Ge, T., Giddaluru, S., Goldman, A.L., Green, M.J., Groenewold, N.A., Grotegerd, D., Gurholt, T.P., Gutman, B.A., Hansell, N.K., Harris, M.A., Harrison, M.B., Haswell, C.C., Hauser, M., Herms, S., Heslenfeld, D.J., Ho, N.F., Hoehn, D., Hoffmann, P., Holleran, L., Hoogman, M., Hottenga, J.J., Ikeda, M., Janowitz, D., Jansen, I.E., Jia, T., Jockwitz, C., Kanai, R., Karama, S., Kasperaviciute, D., Kaufmann, T., Kelly, S., Kikuchi, M., Klein, M., Knapp, M., Knodt, A.R., Krämer, B., Lam, M., Lancaster, T.M., Lee, P.H., Lett, T.A., Lewis, L.B., Lopes-Cendes, I., Luciano, M., Macciardi, F., Marquand, A.F., Mathias, S.R., Melzer, T.R., Milaneschi, Y., Mirza-Schreiber, N., Moreira, J.C.V., Mühleisen, T.W., Müller-Myhsok, B., Najt, P., Nakahara, S., Nho, K., Olde Loohuis, L.M., Orfanos, D.P., Pearson, J.F., Pitcher, T.L., Pütz, B., Quidé, Y., Ragothaman, A., Rashid, F.M., Reay, W.R., Redlich, R., Reinbold, C.S., Repple, J., Richard, G., Riedel, B.C., Risacher, S.L., Rocha, C.S., Mota, N.R., Salminen, L., Saremi, A., Saykin, A.J., Schlag, F., Schmaal, L., Schofield, P.R., Secolin, R., Shapland, C.Y., Shen, L., Shin, J., Shumskaya, E., Sønderby, I.E., Sprooten, E., Tansey, K.E., Teumer, A., Thalamuthu, A., Tordesillas-Gutiérrez, D., Turner, J.A., Uhlmann, A., Vallerga, C.L., van der Meer, D., van Donkelaar, M.M.J., van Eijk, L., van Erp, T.G.M., van Haren, N.E.M., van Rooij, D., van Tol, M.J., Veldink, J.H., Verhoef, E., Walton, E., Wang, M., Wang, Y., Wardlaw, J.M., Wen, W., Westlye, L.T., Whelan, C.D., Witt, S.H., Wittfeld, K., Wolf, C., Wolfers, T., Wu, J.Q., Yasuda, C.L., Zaremba, D., Zhang, Z., Zwiers, M.P., Artiges, E., Assareh, A.A., Ayesa-Arriola, R., Belger, A., Brandt, C.L., Brown, G.G., Cichon, S., Curran, J.E., Davies, G.E., Degenhardt, F., Dennis, M.F., Dietsche, B., Djurovic, S., Doherty, C.P., Espiritu, R., Garijo, D., Gil, Y., Gowland, P.A., Green, R.C., Häusler, A.N., Heindel, W., Ho, B.C., Hoffmann, W.U., Holsboer, F., Homuth, G., Hosten, N., Jack, C.R., Jang, M.H., Jansen, A., Kimbrel, N.A., Kolskår, K., Koops, S., Krug, A., Lim, K.O., Luykx, J.J., Mathalon, D.H., Mather, K.A., Mattay, V.S., Matthews, S., van Son, J.M., McEwen, S.C., Melle, I., Morris, D.W., Mueller, B.A., Nauck, M., Nordvik, J.E., Nöthen, M.M., O’Leary, D.S., Opel, N., Martinot, M.L.P., Bruce Pike, G., Preda, A., Quinlan, E.B., Rasser, P.E., Ratnakar, V., Reppermund, S., Steen, V.M., Tooney, P.A., Torres, F.R., Veltman, D.J., Voyvodic, J.T., Whelan, R., White, T., Yamamori, H., Adams, H.H.H., Bis, J.C., Debette, S., Decarli, C., Fornage, M., Gudnason, V., Hofer, E., Arfan Ikram, M., Launer, L., Longstreth, W.T., Lopez, O.L., Mazoyer, B., Mosley, T.H., Roshchupkin, G. V., Satizabal, C.L., Schmidt, R., Seshadri, S., Yang, Q., Alvim, M.K.M., Ames, D., Anderson, T.J., Andreassen, O.A., Arias-Vasquez, A., Bastin, M.E., Baune, B.T., Beckham, J.C., Blangero, J., Boomsma, D.I., Brodaty, H., Brunner, H.G., Buckner, R.L., Buitelaar, J.K., Bustillo, J.R., Cahn, W., Cairns, M.J., Calhoun, V., Carr, V.J., Caseras, X., Caspers, S., Cavalleri, G.L., Cendes, F., Corvin, A., Crespo-Facorro, B., Dalrymple-Alford, J.C., Dannlowski, U., de Geus, E.J.C., Deary, I.J., Delanty, N., Depondt, C., Desrivières, S., Donohoe, G., Espeseth, T., Fernández, G., Fisher, S.E., Flor, H., Forstner, A.J., Francks, C., Franke, B., Glahn, D.C., Gollub, R.L., Grabe, H.J., Gruber, O., Håberg, A.K., Hariri, A.R., Hartman, C.A., Hashimoto, R., Heinz, A., Henskens, F.A., Hillegers, M.H.J., Hoekstra, P.J., Holmes, A.J., Elliot Hong, L., Hopkins, W.D., Hulshoff Pol, H.E., Jernigan, T.L., Jönsson, E.G., Kahn, R.S., Kennedy, M.A., Kircher, T.T.J., Kochunov, P., Kwok, J.B.J., Le Hellard, S., Loughland, C.M., Martin, N.G., Martinot, J.L., McDonald, C., McMahon, K.L., Meyer-Lindenberg, A., Michie, P.T., Morey, R.A., Mowry, B., Nyberg, L., Oosterlaan, J., Ophoff, R.A., Pantelis, C., Paus, T., Pausova, Z., Penninx, B.W.J.H., Polderman, T.J.C., Posthuma, D., Rietschel, M., Roffman, J.L., Rowland, L.M., Sachdev, P.S., Sämann, P.G., Schall, U., Schumann, G., Scott, R.J., Sim, K., Sisodiya, S.M., Smoller, J.W., Sommer, I.E., Pourcain, B.S., Stein, D.J., Toga, A.W., Trollor, J.N., van der Wee, N.J.A., van’t Ent, D., Völzke, H., Walter, H., Weber, B., Weinberger, D.R., Wright, M.J., Zhou, J., Stein, J.L., Thompson, P.M., Medland, S.E., 2020. The genetic architecture of the human cerebral cortex. Science (1979) 367. 10.1126/science.aay6690

Hallgrimsson, B., Percival, C.J., Green, R., Young, N.M., Mio, W., Marcucio, R., 2015. Morphometrics, 3D Imaging, and Craniofacial Development. Curr Top Dev Biol 115, 561–597. 10.1016/bs.ctdb.2015.09.003

Hata, K., Iida, J., Iwasaka, H., Negoro, H., Kishimoto, T., 2003. Association between minor physical anomalies and lateral ventricular enlargement in childhood and adolescent onset schizophrenia. Acta Psychiatr Scand 108, 147–151. 10.1034/j.1600-0447.2003.00116.x

Hennessy, R.J., Baldwin, P.A., Browne, D.J., Kinsella, A., Waddington, J.L., 2010. Frontonasal dysmorphology in bipolar disorder by 3D laser surface imaging and geometric morphometrics : Comparisons with schizophrenia. Schizophr Res 122, 63–71. 10.1016/j.schres.2010.05.001

Hennessy, R.J., Baldwin, P.A., Browne, D.J., Kinsella, A., Waddington, J.L., 2007. Three-Dimensional Laser Surface Imaging and Geometric Morphometrics Resolve Frontonasal Dysmorphology in Schizophrenia. Biol Psychiatry 61, 1187–1194. 10.1016/j.biopsych.2006.08.045

Hennessy, R.J., Kinsella, A., Waddington, J.L., 2002. 3D laser surface scanning and geometric morphometric analysis of craniofacial shape as an index of cerebro-craniofacial morphogenesis: Initial application to sexual dimorphism. Biol Psychiatry 51, 507–514. 10.1016/S0006-3223(01)01327-0

Hennessy, R.J., Lane, A., Kinsella, A., Larkin, C., Callaghan, E.O., Waddington, J.L., 2004. 3D morphometrics of craniofacial dysmorphology reveals sex-specific asymmetries in schizophrenia. Schizophr Res 67, 261–268. 10.1016/j.schres.2003.08.003

Henriksson, K.M., Wickstrom, K., Maltesson, N., Ericsson, A., Karlsson, J., Lindgren, F., Astrom, K., McNeil, T.F., Agartz, I., 2006. A pilot study of facial, cranial and brain MRI morphometry in men with schizophrenia : Part 2. Psychiatry Res Neuroimaging 147, 187–195. 10.1016/j.pscychresns.2006.03.004

Hibar, D.P., Westlye, L.T., Doan, N.T., Jahanshad, N., Cheung, J.W., Ching, C.R.K., Versace, A., Bilderbeck, A.C., Uhlmann, A., Mwangi, B., Krämer, B., Overs, B., Hartberg, C.B., Abe, C., Dima, D., Grotegerd, D., Sprooten, E., Ben, E., Jimenez, E., Howells, F.M., Delvecchio, G., Temmingh, H., Starke, J., Almeida, J.R.C., Goikolea, J.M., Houenou, J., Beard, L.M., Rauer, L., Abramovic, L., Bonnin, M., Ponteduro, M.F., Keil, M., Rive, M.M., Yao, N., Yalin, N., Najt, P., Rosa, P.G., Redlich, R., Trost, S., Hagenaars, S., Fears, S.C., Alonso-Lana, S., Van Erp, T.G.M., Nickson, T., Chaim-Avancini, T.M., Meier, T.B., Elvsashagen, T., Haukvik, U.K., Lee, W.H., Schene, A.H., Lloyd, A.J., Young, A.H., Nugent, A., Dale, A.M., Pfennig, A., McIntosh, A.M., Lafer, B., Baune, B.T., Ekman, C.J., Zarate, C.A., Bearden, C.E., Henry, C., Simhandl, C., McDonald, C., Bourne, C., Stein, D.J., Wolf, D.H., Cannon, D.M., Glahn, D.C., Veltman, D.J., Pomarol-Clotet, E., Vieta, E., Canales-Rodriguez, E.J., Nery, F.G., Duran, F.L.S., Busatto, G.F., Roberts, G., Pearlson, G.D., Goodwin, G.M., Kugel, H., Whalley, H.C., Ruhe, H.G., Soares, J.C., Fullerton, J.M., Rybakowski, J.K., Savitz, J., Chaim, K.T., Fatjó-Vilas, M., Soeiro-De-Souza, M.G., Boks, M.P., Zanetti, M. V., Otaduy, M.C.G., Schaufelberger, M.S., Alda, M., Ingvar, M., Phillips, M.L., Kempton, M.J., Bauer, M., Landén, M., Lawrence, N.S., Van Haren, N.E.M., Horn, N.R., Freimer, N.B., Gruber, O., Schofield, P.R., Mitchell, P.B., Kahn, R.S., Lenroot, R., Machado-Vieira, R., Ophoff, R.A., Sarró, S., Frangou, S., Satterthwaite, T.D., Hajek, T., Dannlowski, U., Malt, U.F., Arolt, V., Gattaz, W.F., Drevets, W.C., Caseras, X., Agartz, I., Thompson, P.M., Andreassen, O.A., 2018. Cortical abnormalities in bipolar disorder: An MRI analysis of 6503 individuals from the ENIGMA Bipolar Disorder Working Group. Mol Psychiatry 23, 932–942. 10.1038/mp.2017.73

Howes, O.D., Cummings, C., Chapman, G.E., Shatalina, E., 2023. Neuroimaging in schizophrenia: an overview of findings and their implications for synaptic changes. Neuropsychopharmacology 48, 151–167. 10.1038/s41386-022-01426-x

Jauhar, S., Johnstone, M., McKenna, P.J., 2022. Schizophrenia. The Lancet 399, 473–486. 10.1016/S0140-6736(21)01730-X

Jha, S.C., Xia, K., Schmitt, J.E., Ahn, M., Girault, J.B., Murphy, V.A., Li, G., Wang, L., Shen, D., Zou, F., Zhu, H., Styner, M., Knickmeyer, R.C., Gilmore, J.H., 2018. Genetic influences on neonatal cortical thickness and surface area. Hum Brain Mapp 39, 4998–5013. 10.1002/hbm.24340

Karantonis, J.A., Rossell, S.L., Carruthers, S.P., Sumner, P., Hughes, M., Green, M.J., Pantelis, C., Burdick, K.E., Cropley, V., Van Rheenen, T.E., 2020. Cognitive validation of cross-diagnostic cognitive subgroups on the schizophrenia-bipolar spectrum. J Affect Disord 266, 710–721. 10.1016/j.jad.2020.01.123

Katina, S., Kelly, B.D., Rojas, M.A., Sukno, F.M., McDermott, A., Hennessy, R.J., Lane, A., Whelan, P.F., Bowman, A.W., Waddington, J.L., 2020. Refining the resolution of craniofacial dysmorphology in bipolar disorder as an index of brain dysmorphogenesis. Psychiatry Res 291, 113243. 10.1016/j.psychres.2020.113243

Kelly, B.D., McNeil, T.F., Lane, A., Henriksson, K.M., Kinsella, A., Agartz, I., 2005. Is craniofacial dysmorpholgy correlated with structural brain anomalies in schizophrenia? Schizophr Res 80, 349–355. 10.1016/j.schres.2005.07.036

Kesterke, M.J., Raffensperger, Z.D., Heike, C.L., Cunningham, M.L., Hecht, J.T., Kau, C.H., Nidey, N.L., Moreno, L.M., Wehby, G.L., Marazita, M.L., Weinberg, S.M., 2016. Using the 3D Facial Norms Database to investigate craniofacial sexual dimorphism in healthy children, adolescents, and adults. Biol Sex Differ 7. 10.1186/s13293-016-0076-8

Klingenberg, C.P., 2016. Size, shape, and form: concepts of allometry in geometric morphometrics. Dev Genes Evol 226, 113–137. 10.1007/s00427-016-0539-2

Klingenberg, C.P., 2011. MORPHO J : an integrated software package for geometric morphometrics. Mol Ecol Resour 11, 353–357. 10.1111/j.1755-0998.2010.02924.x

Kloiber, S., Rosenblat, J.D., Husain, M.I., Ortiz, A., Berk, M., Quevedo, J., Vieta, E., Maes, M., Birmaher, B., Soares, J.C., Carvalho, A.F., 2020. Neurodevelopmental pathways in bipolar disorder. Neurosci Biobehav Rev 112, 213–226. 10.1016/j.neubiorev.2020.02.005

Lane, A., Kinsella, A., Murphy, P., Byrne, M., Keenan, J., Colgan, K., Cassidy, B., Sheppard, N., Horgan, R., Waddington, J.L., Larkin, C., O’Callaghan, E., 1997. The anthropometric assessment of dysmorphic features in schizophrenia as an index of its developmental origins. Psychol Med 27, 1155–1164. 10.1017/S0033291797005503

Larson, J.R., Manyama, M.F., Cole, J.B., Gonzalez, P.N., Percival, C.J., Liberton, D.K., Ferrara, T.M., Riccardi, S.L., Kimwaga, E.A., Mathayo, J., Spitzmacher, J.A., Rolian, C., Jamniczky, H.A., Weinberg, S.M., Roseman, C.C., Klein, O., Lukowiak, K., Spritz, R.A., Hallgrimsson, B., 2018. Body size and allometric variation in facial shape in children. Am J Phys Anthropol 165, 327–342. 10.1002/ajpa.23356

Lawrie, S.M., Byrne, M., Miller, P., Hodges, A., Clafferty, R.A., Cunningham Owens, D.G., Johnstone, E.C., 2001. Neurodevelopmental indices and the development of psychotic symptoms in subjects at high risk of schizophrenia. British Journal of Psychiatry 178, 524–530. 10.1192/bjp.178.6.524

Lele, S., Richtsmeier, J.T., 2001. An Invariant Approach to Statistical Analysis of Shapes, Chapman \& Hall/Crc Interdisciplinary Statistics. CRC Press.

Liu, S., Rao, S., Xu, Y., Li, J., Huang, H., Zhang, X., Fu, H., Wang, Q., Cao, H., Baranova, A., Jin, C., Zhang, F., 2020. Identifying common genome-wide risk genes for major psychiatric traits. Hum Genet 139, 185–198. 10.1007/s00439-019-02096-4

Marcucio, Ralph Hallgrimsson, B., Young, Nathan., 2015. Facial Morphogenesis: Physical and Molecular Interactions Between the Brain and the Face. Curr Top Dev Biol 115, 229–319. 10.1016/bs.ctdb.2015.09.001.

Mitteroecker, P., Gunz, P., 2009. Advances in Geometric morphometrics. Evol Biol 36, 235–247. 10.1007/s11692-009-9055-x

Mitteroecker, P., Schaefer, K., 2022. Thirty years of geometric morphometrics: Achievements, challenges, and the ongoing quest for biological meaningfulness. American Journal of Biological Anthropology 178, 181–210. 10.1002/ajpa.24531

Naqvi, S., Sleyp, Y., Hoskens, H., Indencleef, K., Spence, J.P., Bruffaerts, R., Radwan, A., Eller, R.J., Richmond, S., Shriver, M.D., Shaffer, J.R., Weinberg, S.M., Walsh, S., Thompson, J., Pritchard, J.K., Sunaert, S., Peeters, H., Wysocka, J., Claes, P., 2021. Shared heritability of human face and brain shape. Nat Genet 53, 830–839. 10.1038/s41588-021-00827-w

Nelson, H.E., Willison, J., 1991. National adult reading test (NART). Nfer-Nelson Windsor.

Norbom, L.B., Ferschmann, L., Parker, N., Agartz, I., Andreassen, O.A., Paus, T., Westlye, L.T., Tamnes, C.K., 2021. New insights into the dynamic development of the cerebral cortex in childhood and adolescence: Integrating macro- and microstructural MRI findings. Prog Neurobiol 204. 10.1016/j.pneurobio.2021.102109

O’Callaghan, E., Buckley, P., Madigan, C., Redmond, O., Stack, J.P., Kinsella, A., Larkin, C., Ennis, J.T., Waddington, J.L., 1995. The relationship of minor physical anomalies and other putative indices of developmental disturbance in schizophrenia to abnormalities of cerebral structure on magnetic resonance imaging. Biol Psychiatry 38, 516–524. 10.1016/0006-3223(94)00381-C

Owen, M.J., O’Donovan, M.C., 2017. Schizophrenia and the neurodevelopmental continuum:evidence from genomics. World Psychiatry 16, 227–235. 10.1002/wps.20440

Patel, Y., Shin, J., Abé, C., Agartz, I., Alloza, C., Alnæs, D., Ambrogi, S., Antonucci, L.A., Arango, C., Arolt, V., Auzias, G., Ayesa-Arriola, R., Banaj, N., Banaschewski, T., Bandeira, C., Başgöze, Z., Cupertino, R.B., Bau, C.H.D., Bauer, J., Baumeister, S., Bernardoni, F., Bertolino, A., Bonnin, C. del M., Brandeis, D., Brem, S., Bruggemann, J., Bülow, R., Bustillo, J.R., Calderoni, S., Calvo, R., Canales-Rodríguez, E.J., Cannon, D.M., Carmona, S., Carr, V.J., Catts, S. V., Chenji, S., Chew, Q.H., Coghill, D., Connolly, C.G., Conzelmann, A., Craven, A.R., Crespo-Facorro, B., Cullen, K., Dahl, A., Dannlowski, U., Davey, C.G., Deruelle, C., Díaz-Caneja, C.M., Dohm, K., Ehrlich, S., Epstein, J., Erwin-Grabner, T., Eyler, L.T., Fedor, J., Fitzgerald, J., Foran, W., Ford, J.M., Fortea, L., Fuentes-Claramonte, P., Fullerton, J., Furlong, L., Gallagher, L., Gao, B., Gao, S., Goikolea, J.M., Gotlib, I., Goya-Maldonado, R., Grabe, H.J., Green, M., Grevet, E.H., Groenewold, N.A., Grotegerd, D., Gruber, O., Haavik, J., Hahn, T., Harrison, B.J., Heindel, W., Henskens, F., Heslenfeld, D.J., Hilland, E., Hoekstra, P.J., Hohmann, S., Holz, N., Howells, F.M., Ipser, J.C., Jahanshad, N., Jakobi, B., Jansen, A., Janssen, J., Jonassen, R., Kaiser, A., Kaleda, V., Karantonis, J., King, J.A., Kircher, T., Kochunov, P., Koopowitz, S.M., Landén, M., Landrø, N.I., Lawrie, S., Lebedeva, I., Luna, B., Lundervold, A.J., MacMaster, F.P., Maglanoc, L.A., Mathalon, D.H., McDonald, C., McIntosh, A., Meinert, S., Michie, P.T., Mitchell, P., Moreno- Alcázar, A., Mowry, B., Muratori, F., Nabulsi, L., Nenadić, I., O’Gorman Tuura, R., Oosterlaan, J., Overs, B., Pantelis, C., Parellada, M., Pariente, J.C., Pauli, P., Pergola, G., Piarulli, F.M., Picon, F., Piras, F., Pomarol-Clotet, E., Pretus, C., Quidé, Y., Radua, J., Ramos-Quiroga, J.A., Rasser, P.E., Reif, A., Retico, A., Roberts, G., Rossell, S., Rovaris, D.L., Rubia, K., Sacchet, M., Salavert, J., Salvador, R., Sarró, S., Sawa, A., Schall, U., Scott, R., Selvaggi, P., Silk, T., Sim, K., Skoch, A., Spalletta, G., Spaniel, F., Stein, D.J., Steinsträter, O., Stolicyn, A., Takayanagi, Y., Tamm, L., Tavares, M., Teumer, A., Thiel, K., Thomopoulos, S.I., Tomecek, D., Tomyshev, A.S., Tordesillas-Gutiérrez, D., Tosetti, M., Uhlmann, A., Van Rheenen, T., Vazquez-Bourgón, J., Vernooij, M.W., Vieta, E., Vilarroya, O., Weickert, C., Weickert, T., Westlye, L.T., Whalley, H., Willinger, D., Winter, A., Wittfeld, K., Yang, T.T., Yoncheva, Y., Zijlmans, J.L., Hoogman, M., Franke, B., van Rooij, D., Buitelaar, J., Ching, C.R.K., Andreassen, O.A., Pozzi, E., Veltman, D., Schmaal, L., van Erp, T.G.M., Turner, J., Castellanos, F.X., Pausova, Z., Thompson, P., Paus, T., 2022. Virtual Ontogeny of Cortical Growth Preceding Mental Illness. Biol Psychiatry 92, 299–313. 10.1016/j.biopsych.2022.02.959

Radua, J., Murray, R.M., Ramella-Cravaro, V., Amir, T., McGuire, P., Yenn Thoo, H., Ioannidis, J.P.A., Phiphopthatsanee, N., Reichenberg, A., Morgan, C., Davies, C., Oliver, D., Fusar-Poli, P., 2018. What causes psychosis? An umbrella review of risk and protective factors. World Psychiatry 49–66. 10.1002/wps.20490

Sandman, C.A., Glynn, L.M., Davis, E.P., 2013. Is there a viability-vulnerability tradeoff? Sex differences in fetal programming. J Psychosom Res 75, 327–335. 10.1016/j.jpsychores.2013.07.009

Starbuck, J.M., Llambrich, S., Gonzàlez, R., Albaigès, J., Sarlé, A., Wouters, J., González, A., Sevillano, X., Sharpe, J., De La Torre, R., Dierssen, M., Vande Velde, G., Martínez-Abadías, N., 2021. Green tea extracts containing epigallocatechin-3-gallate modulate facial development in Down syndrome. Sci Rep 11. 10.1038/s41598-021-83757-1

The MathWorks Inc, 2022. MATLAB.

van Erp, T.G.M., Walton, E., Hibar, D.P., Schmaal, L., Jiang, W., Glahn, D.C., Pearlson, G.D., Yao, N., Fukunaga, M., Hashimoto, R., Okada, N., Yamamori, H., Bustillo, J.R., Clark, V.P., Agartz, I., Mueller, B.A., Cahn, W., de Zwarte, S.M.C., Hulshoff Pol, H.E., Kahn, R.S., Ophoff, R.A., van Haren, N.E.M., Andreassen, O.A., Dale, A.M., Doan, N.T., Gurholt, T.P., Hartberg, C.B., Haukvik, U.K., Jørgensen, K.N., Lagerberg, T. V., Melle, I., Westlye, L.T., Gruber, O., Kraemer, B., Richter, A., Zilles, D., Calhoun, V.D., Crespo-Facorro, B., Roiz-Santiañez, R., Tordesillas-Gutiérrez, D., Loughland, C., Carr, V.J., Catts, S., Cropley, V.L., Fullerton, J.M., Green, M.J., Henskens, F.A., Jablensky, A., Lenroot, R.K., Mowry, B.J., Michie, P.T., Pantelis, C., Quidé, Y., Schall, U., Scott, R.J., Cairns, M.J., Seal, M., Tooney, P.A., Rasser, P.E., Cooper, G., Shannon Weickert, C., Weickert, T.W., Morris, D.W., Hong, E., Kochunov, P., Beard, L.M., Gur, R.E., Gur, R.C., Satterthwaite, T.D., Wolf, D.H., Belger, A., Brown, G.G., Ford, J.M., Macciardi, F., Mathalon, D.H., O’Leary, D.S., Potkin, S.G., Preda, A., Voyvodic, J., Lim, K.O., McEwen, S., Yang, F., Tan, Y., Tan, S., Wang, Z., Fan, F., Chen, J., Xiang, H., Tang, S., Guo, H., Wan, P., Wei, D., Bockholt, H.J., Ehrlich, S., Wolthusen, R.P.F., King, M.D., Shoemaker, J.M., Sponheim, S.R., De Haan, L., Koenders, L., Machielsen, M.W., van Amelsvoort, T., Veltman, D.J., Assogna, F., Banaj, N., de Rossi, P., Iorio, M., Piras, F., Spalletta, G., McKenna, P.J., Pomarol-Clotet, E., Salvador, R., Corvin, A., Donohoe, G., Kelly, S., Whelan, C.D., Dickie, E.W., Rotenberg, D., Voineskos, A.N., Ciufolini, S., Radua, J., Dazzan, P., Murray, R., Reis Marques, T., Simmons, A., Borgwardt, S., Egloff, L., Harrisberger, F., Riecher-Rössler, A., Smieskova, R., Alpert, K.I., Wang, L., Jönsson, E.G., Koops, S., Sommer, I.E.C., Bertolino, A., Bonvino, A., Di Giorgio, A., Neilson, E., Mayer, A.R., Stephen, J.M., Kwon, J.S., Yun, J.Y., Cannon, D.M., McDonald, C., Lebedeva, I., Tomyshev, A.S., Akhadov, T., Kaleda, V., Fatouros-Bergman, H., Flyckt, L., Farde, L., Flyckt, L., Engberg, G., Erhardt, S., Fatouros-Bergman, H., Cervenka, S., Schwieler, L., Piehl, F., Agartz, I., Collste, K., Victorsson, P., Malmqvist, A., Hedberg, M., Orhan, F., Busatto, G.F., Rosa, P.G.P., Serpa, M.H., Zanetti, M. V., Hoschl, C., Skoch, A., Spaniel, F., Tomecek, D., Hagenaars, S.P., McIntosh, A.M., Whalley, H.C., Lawrie, S.M., Knöchel, C., Oertel-Knöchel, V., Stäblein, M., Howells, F.M., Stein, D.J., Temmingh, H.S., Uhlmann, A., Lopez-Jaramillo, C., Dima, D., McMahon, A., Faskowitz, J.I., Gutman, B.A., Jahanshad, N., Thompson, P.M., Turner, J.A., 2018. Cortical Brain Abnormalities in 4474 Individuals With Schizophrenia and 5098 Control Subjects via the Enhancing Neuro Imaging Genetics Through Meta Analysis (ENIGMA) Consortium. Biol Psychiatry 84, 644–654. 10.1016/j.biopsych.2018.04.023

Waddington, J.L., Lane, A., Scully, P., Meagher, D., Quinn, J., Larkin, C., O’Callaghan, E., 1999. Early cerebro-craniofacial dysmorphogenesis in schizophrenia: A lifetime trajectory model from neurodevelopmental basis to “neuroprogressive” process. J Psychiatr Res 33, 477–489. 10.1016/S0022-3956(99)00024-2

Weinberger, D.R., 1995. From neuropathology to neurodevelopment. The Lancet 346, 552–557.

Weinberger, D.R., Levitt, P., 2011. Neurodevelopmental origins of schizophrenia, in: Weinberger, D.R., Harrisson, P. (Eds.), Schizophrenia. Wiley-Blackwell, Oxford, pp. 393–412.

Wierenga, L.M., Langen, M., Oranje, B., Durston, S., 2014. Unique developmental trajectories of cortical thickness and surface area. Neuroimage 87, 120–126. 10.1016/j.neuroimage.2013.11.010

Winkler, A.M., Kochunov, P., Blangero, J., Almasy, L., Zilles, K., Fox, P.T., Duggirala, R., Glahn, D.C., 2010. Cortical thickness or grey matter volume? The importance of selecting the phenotype for imaging genetics studies. Neuroimage 53, 1135–46. 10.1016/j.neuroimage.2009.12.028

