## Supplementary Materials 2 for "Face-brain correlates as potential sex-specific biomarkers for schizophrenia and bipolar disorder"

**Method 1: Euclidean Distance Matrix Analysis and Facial Difference Score**

The Euclidean Distance Matrix Analysis (EDMA) and the computation of the Facial Difference Score (FDS) starts with a population divided into two groups: healthy controls and patients (either with bipolar disorder or schizophrenia), which will be referred hereafter as $HG$ and $PG$. Without loss of generality, we consider that the number of individuals in each group is $n_{H}$ and $n_{P}$, respectively, and that a total of $L$ landmarks have been manually recorded on the 3D facial reconstruction of each individual. As EDMA represents each individual as a matrix of linear distances between all possible pairs of landmarks, there exist $K=\left( \begin{aligned} L \\ 2 \end{aligned} \right)=\frac{L!}{\left( L-2 \right)!2!}$ distances between all possible pairs of landmarks, which we refer to as interlandmark *distances*. In our analysis, as $L=20$, $K=190$. The method to perform EDMA and compute FDS followed these steps:

1. We computed the $K$ interlandmark distances on the $n_{H}$ individuals in $HG$. Then, we computed the average value of each interlandmark distance ($\mu_{HG}^{i}$, with $i=\left\{ 1, \ldots, K \right\}$)) and their joint covariance matrix $C_{HG}=\left[ \begin{matrix} \sigma_{HG}^{11} & \cdots& \sigma_{HG}^{1K} \\ \vdots& \ddots& \vdots\\ \sigma_{HG}^{K1} & \cdots& \sigma_{HG}^{KK} \end{matrix} \right]$ (where $\sigma_{HG}^{ij}$ stands for the covariance between the interlandmark distances $i$ and $j$), and use these to model a multivariate normal distribution $N\left( \left[ \mu_{HG}^{1} \mu_{HG}^{2}\ldots\mu_{HG}^{K} \right], C_{HG} \right)$ of the interlandmark distances of the healthy control group $HG$.

2. We repeated step 1 but now on the patients group $PG$. As a result, we obtained the average value of the $K$ interlandmark distances and their joint covariance matrix, $\mu_{PG}^{i}$ (with $i=\left\{ 1, \ldots, K \right\}$) and $C_{PG}=\left[ \begin{matrix} \sigma_{PG}^{11} & \cdots& \sigma_{PG}^{1K} \\ \vdots& \ddots& \vdots\\ \sigma_{PG}^{K1} & \cdots& \sigma_{PG}^{KK} \end{matrix} \right]$, and used these matrices to model a multivariate normal distribution $N\left( \left[ \mu_{PG}^{1} \mu_{PG}^{2}\ldots\mu_{PG}^{K} \right], C_{PG} \right)$ of the interlandmark distances of the patients group $PG$.

Next, we performed an iterative procedure to obtain the FDS. Steps 3 to 5 were repeated 5,000^[1]^ times:

3. Used the aforementioned multivariate normal distributions to randomly generate $n_{H}$ and $n_{P}$ artificial individuals. We referred to these as the artificial healthy controls and patients groups, $AHG$ and $APG$, respectively.

4. On each artificial group, we computed the average value of each interlandmark distance. For instance, the average value between landmarks $L_{a}$ and $L_{b}$ would be $\mu_{AHG}^{L_{a}\leftrightarrow L_{b}}$ and $\mu_{APG}^{L_{a}\leftrightarrow L_{b}}$ for the artificial healthy controls and patients groups, respectively.

5. For each of the $K$ interlandmark distances, we computed the difference between both averages: $\mu_{AHG}^{L_{a}\leftrightarrow L_{b}}-\mu_{APG}^{L_{a}\leftrightarrow L_{b}}$, and stored the differences.

6. Subsequently, we analyzed which interlandmark distances were significantly different between the 5,000 random $AHG$ and $APG$ groups. To that end, for each interlandmark distance, we ranked the 5,000 differences in increasing order, and analyzed both ends of the 90% confidence interval (CI) to determine whether that particular interlandmark distance was significantly different between the $HG$ and $PG$ groups:

i) if one end was positive and the other negative, the null difference was included in the CI, suggesting that the interlandmark distance was not significantly different between healthy controls and patients

ii) if both ends of the CI were positive, the interlandmark distancebetween landmarks $L_{a}$ and $L_{b}$ was considered as significantly larger in healthy controls in patients comparison to patients.

iii) if both ends of the CI were negative, the interlandmark distance between landmarks $L_{a}$ and $L_{b}$ was considered as significantly smaller in healthy controls than in patients.

7. When this analysis was completed on all the interlandmark distances, we computed the FDS as the percentage of the $K$ interlandmark distances that were significantly different between both groups.

8. In addition to FDS, we also computed the extent of differences in the interlandmark distances that were found to be significantly distinct between the two groups. To compute the relative difference by taking the average of the confidence interval endpoints and dividing it by the average value of that specific interlandmark distance within the healthy control population.

**Method 2: EDMA iterative bootstrapping simulations**

To determine if the FDS between the healthy control and patient groups significantly differed from the FDS that could occur by chance solely among healthy controls, we employed an iterative bootstrapping method based on EDMA.

Steps 1 to 6 were executed iteratively 200^[2]^ times:

1. We randomly subsampled the healthy controls and patients groups $HG$ and $PG$, creating two samples of size $s$ (with $s<n_{H}$ and $s<n_{P}$), which were defined as follows:

- Healthy controls sample ($HS$): formed by $s$ randomly chosen controls individuals from $HG$.
- Mixed sample ($MS$): formed by $n$ healthy controls (randomly chosen from $HG$) and $s-n$ patients (randomly chosen from $PG$).

2. We computed the $K$ interlandmark distances on the $s$ individuals in $HS$. Then, we computed the average value of each interlandmark distance ($\mu_{HS}^{i}$, with $i=\left\{ 1, \ldots, K \right\}$)) and their joint covariance matrix $C_{HS}=\left[ \begin{matrix} \sigma_{HS}^{11} & \cdots& \sigma_{HS}^{1K} \\ \vdots& \ddots& \vdots\\ \sigma_{HS}^{K1} & \cdots& \sigma_{HS}^{KK} \end{matrix} \right]$ , where $\sigma_{HG}^{ij}$ stands for the covariance between the interlandmark distances $i$ and $j$), and used these to model a multivariate normal distribution $N\left( \left[ \mu_{HS}^{1} \mu_{HS}^{2}\ldots\mu_{HS}^{K} \right], C_{HS} \right)$ of the interlandmark distances of the healthy controls sample $HS$.

3. We computed the $K$ interlandmark distances on the $s$ individuals in $MS$. Then, we computed the average value of each interlandmark distance ($\mu_{MS}^{i}$, with $i=\left\{ 1, \ldots, K \right\}$)) and their joint covariance matrix $C_{MS}=\left[ \begin{matrix} \sigma_{MS}^{11} & \cdots& \sigma_{MS}^{1K} \\ \vdots& \ddots& \vdots\\ \sigma_{MS}^{K1} & \cdots& \sigma_{MS}^{KK} \end{matrix} \right]$ (where $\sigma_{HG}^{ij}$ stands for the covariance between the interlandmark distances $i$ and $j$), and used these to model a multivariate normal distribution $N\left( \left[ \mu_{MS}^{1} \mu_{MS}^{2}\ldots\mu_{MS}^{K} \right], C_{MS} \right)$ of the interlandmark distances of the mixed sample $MS$.

4. Next, steps 4.1 to 4.4 were executed 5,000 times:

4.1. We used the aforementioned multivariate normal distributions to randomly generate $s$ artificial individuals of each class. We referred to these as the artificial healthy controls and mixed samples, $AHS$ and $AMS$, respectively.

4.2. On each artificial sample, we computed the average value of each interlandmark distance. For instance, the average value between landmarks $L_{a}$ and $L_{b}$ would be $\mu_{AHS}^{L_{a}\leftrightarrow L_{b}}$ and $\mu_{AMS}^{L_{a}\leftrightarrow L_{b}}$ for the artificial healthy controls and mixed samples, respectively.

4.3. For each of the $K$ interlandmark distances, we computed the difference between both averages: $\mu_{AHS}^{L_{a}\leftrightarrow L_{b}}-\mu_{AMS}^{L_{a}\leftrightarrow L_{b}}$.

4.4. We stored the differences between the averages of each one of the $K$ interlandmark distances.

5. To determine whether that particular interlandmark distance was significantly different between the $HS$ and $MS$ samples, we repeated step 6 from Method 1.

6. To measure the morphological difference between both samples, we computed the percentage of the $K$ interlandmark distances that were significantly different between them and stored these values.

7. After the 200 executions of steps 1 to 6, we created a histogram to represent the percentage of the $K$ interlandmark distances that were significantly different between the healthy control and the mix samples with the current composition of the latter.

8. We repeated steps 1 to 7, changing progressively the composition of the mix sample $MS$, decreasing the number of healthy controls ($n$) and increasing the number of patients ($s-n$). The goal was that, at the first iteration, the mixed sample $MS$ was composed entirely of randomly chosen healthy controls (i.e. $n=s$), and at the last iteration, the mixed sample $MS$ was composed entirely of randomly chosen patients (i.e. $n=0$).

9. We created a stacked plot of the histograms obtained from each iteration of the method to visualize the trend of the number of significantly different interlandmark distances as the composition of the mixed sample $MS$ evolved from containing only controls to containing only patients.

10. To find the statistical significance of the EDMA iterative bootstrapping, we computed the p-value as the ratio of experiments performed in a $MS$ entirely composed of randomly chosen healthy controls in which the percentage of significantly different interlandmark distances was higher than the FDS value obtained applying Method 1.

1. The number of repetitions (5,000) was set arbitrarily. In any case, the process should be iterated a number of times that ensures that the distributions of the interlandmark distances are sufficiently sampled.
2. Again, this value is arbitrary but should be large enough to ensure the statistical significance of the results.
